# Advanced MRI Prediction Model for Anatomical Site Identification in Uterine Carcinoma: Enhancing Diagnostic Accuracy and Treatment Planning

**DOI:** 10.1101/2024.04.24.24305372

**Authors:** Mahrooz Malek, Moneereh moayeri, Setareh Akhavan, Shahrzad Sheikh Hasani, Fatemeh Nili, Zahra Mahboubi-Fooladi

**Affiliations:** Muskoskletal Imaging Research Research Center (MIRC)Radiology Department, Imam Khomeini Hospital Complex (IKHC), Tehran University of Medical Sciences, Tehran, Iran; Advanced Diagnostic and Interventional Radiology Research Center (ADIR), Radiology Department, Imam Khomeini Hospital Complex (IKHC), Tehran University of Medical Sciences (TUMS), Tehran, Iran; Department of Obstetrics and Gynecology, Vali-Asr Reproductive Health Research Center, Tehran University of Medical Sciences, Tehran, Iran; Department of Oncologic Gynecology, Vali-Asr Hospital, Tehran University of Medical Sciences, Tehran, Iran; Department of Pathology, Cancer Institute, Imam Khomeini Hospital Complex, Tehran, Iran; Department of Radiology, Shahid Beheshti University of Medical Sciences, Tehran, Iran

**Keywords:** oncology, cervical cancer, uterine carcinoma, gynecology

## Abstract

**Introduction:** The uterine carcinoma is the most commonly diagnosed malignancy in female pelvis. Accurate identification of tumor origin is crucial for determining appropriate treatment approaches. This study aims to develop a prediction model using multiple MRI parameters to accurately diagnose uterine cancer with an indistinctive origin and those involving both the endometrium and cervix prior to treatment.

**Material and methods:** This prospective cohort study was conducted from January 2020 to January 2021, and included patients aged 20-80 who were newly diagnosed with uterine carcinoma who underwent MRI and were considered for hysterectomy within 6 months after MRI.

**Results:** In our study, a total of 78 patients with uterine carcinoma were enrolled. the final diagnosis was confirmed as follows: 20 were adenocarcinoma of the cervix, 27 were SCC of the cervix, and 31 were endometrial adenocarcinoma. Certain imaging features were found to be consistent with cervical carcinoma, included parametrial invasion (69.6%), vaginal invasion (66%), stromal invasion (95.7%), and peripheral rim enhancement (68.9%). On post-contrast sequences, cervical cancer appeared hyperintense compared to the myometrium, while endometrial cancer appeared hypointense (96.8%). Endometrial carcinoma was well diagnosed by the presence of an endometrial cavity mass (100%), deep myometrium invasion (>50%) (54.8%), and a greater size in the craniocaudal dimension compared to the transverse dimension (100%).

**Discussion:** The study found that certain morphologic features were reliable indicators for detecting cervical carcinoma, including vaginal, stromal, and parametrial invasions, the presence of hypervascularity and peripheral rim enhancement. On the other hand, myometrial invasion and the presence of a mass in the endometrial cavity were significantly higher in endometrial carcinoma.

## Introduction

The uterine carcinoma is the most commonly diagnosed malignancy in female pelvis in the United States and can originate from either cervix or endometrium. Accurately identifying the anatomical site of tumor origin is crucial, as tumors located in the cervix exhibit different biological and clinical behavior compared to those located in the endometrium, requiring different treatment approaches (1, 2, 3).

Simple hysterectomy with bilateral salpingo-oophorectomy is recommended for both staging and treatment of endometrial carcinoma, whereas the preferred approach for cervical carcinoma is assorted based on histologic examination. Stage IB1-IIA1 cervical carcinoma is typically treated with radical hysterectomy and pelvic lymphadenectomy, while stage IIB or higher is managed with chemoradiation (3, 4, 5, 6). An inaccurate diagnosis may lead to improper treatment selection (3).

Although the origin of most of the newly diagnosed uterine cancers can be easily determined, clinicians may encounter difficulties in discriminating the origin of tumor due to inconclusive biopsy results (mixed type or unusual histologic findings, absence of precursor lesions, or inadequate samples) (3).

Several studies have demonstrated the advantages of MRI in determining the anatomical origin of tumors, but most of them rely on subjective assessments. Additionally, the staging of uterine cancers according to the International Federation of Gynecology and Obstetrics (FIGO) has undergone significant revision, highlighting the importance of MRI in depicting morphologic prognostic factors that correlate with tumor grades and proper management. However, there is limited information regarding the utility of MRI as the primary tool in diagnosing and treating large and invasive uterine tumors involving both the endometrium and cervix (7, 8, 9).

The objective of this study is to develop a prediction model that combines multiple parameters, including morphology, tumor site, depth of invasion, and quantitative values of T1 and T2-weighted MRI, as well as contrast enhancement MRI patterns using the myometrium as an internal reference. The aim is to establish MRI as an early and reliable tool for accurately diagnosing uterine cancer with indistinctive origin and those involving both the endometrium and cervix prior to treatment and tissue removal (10).

## Method and material

### Study design and participant patients

This prospective cohort study was approved by the institutional review board committee of Tehran University of Medical Science and adhered to the tenets of the Declaration of Helsinki, written informed consent was obtained from all patients enrolled.

From January 2020 to January 2021, we included patients aged 20-80 who were newly diagnosed with uterine carcinoma (cervical or endometrial), both clinically and initially biopsy-proven. These patients were requested to undergo MRI evaluation as per a routine protocol by a dedicated gynecology oncology team. Additionally, they were considered for hysterectomy within 6 months after MRI.

Exclusion criteria were defined as follows:

1. Contraindications to MRI imaging, such as glomerular filtration rates less than 30 mL/min/1.73m^2, cardiac pacemaker, metal implants, or claustrophobia.
2. Patients with microscopic lesions.
3. Patients who had received hormones, chemotherapy, or radiotherapy.
4. Patients for whom only biopsy sampling with immunohistochemistry (IHC) staining was planned, and not surgery.
5. Unavailability of interpretation of pathology.
6. Patients diagnosed with other types of cancers, apart from adenocarcinoma of the endometrium and both adenocarcinoma and squamous cell carcinoma (SCC) of the cervix.

In this study, a total of 78 patients who were eligible were recruited. After undergoing a hysterectomy, histologic examination was used as the gold standard to determine the definitive diagnosis. Additionally, the MRI images were analyzed to determine if the findings correlated with the final diagnosis.

### MRI protocols

The MRI protocols used in this study involved using a 3T MRI machine (Magnetom Trio, Siemens, Erlangen, Germany) with a four-channel body phased array coil placed over the pelvis. Patients were instructed to fast for three hours prior to the MRI examination. They were also given intra-muscular Hyoscine butyl bromide 30 minutes before the examination as an anti-peristaltic agent.

Routine sequences, including spin echo T1 and T2-weighted sequences, were obtained in the sagittal, axial, and coronal planes to evaluate uterine mass. Additionally, DW-MRI (diffusion-weighted MRI) was conducted using a single-shot spin echo planar sequence with a slice thickness of 8mm and a field of view of 280mm on the axial plane. ADC maps were generated for all DW images. Dynamic contrast-enhanced MR images (DCE-MR) were acquired by administering gadolinium contrast medium at a dose of 0.2mmol/kg. Images were obtained in the sagittal plane with a time resolution of 17 seconds and a scan duration of 3 minutes. This allowed for post-contrast images to be taken every 17 seconds, resulting in a total acquisition time of approximately 3 minutes. The use of contrast enhancement helps improve the accuracy of MRI in evaluating uterine carcinoma.

### MRI Analysis

Two expert gynecological radiologists independently performed the interpretation of MRI images. Utilizing their combined 8 years of experience and documentation from relevant literature, the two readers reached a consensus on the assessment of the parameters listed below. The parameters assessed during the interpretation of the MRI images were as follows:

1. Tumor location: confined as the most prominent site involved by the tumor, either in the cervix or endometrium. However, if the tumor was equally present throughout the cervix and endometrium, the origin was considered to originate from the endometrium.
2. The rim enhancement; whether it was complete or partial
3. Early arterial enhancement: to detect perfusion of the tumor during the early phase (45 seconds) and estimate its enhancement compared to the myometrium. If the contrast was abnormally excessive, resulting in a marked increase in enhancement compared to the myometrium, the tumor was presumed to be hypervascular. Otherwise, it was considered hypovascular.
4. The depth of myometrium invasion is determined by calculating the ratio of the deepest outer tumor margin to the total thickness of the myometrium. If this ratio exceeds 50%, it is classified as deep myometrium invasion.
5. A distended endometrial cavity is defined as an anteroposterior diameter of the fluid-filled cavity exceeding 5mm in the sagittal plane of T2-weighted imaging.
6. The presence of a mass within the endometrial cavity is indicated by the disappearance of normal endometrial hyperintensity on a T2-weighted sequence, accompanied by a soft tissue mass within the uterine cavity.
7. Invasion into the parametrium, vagina, and cervical stroma.
8. For the assessment of tumor size, measurements of both the cranio-caudal and transverse dimensions were performed.

Region-of-interest (ROI) measurements were conducted by manually delineating the largest circular ROI around the tumor, excluding areas with necrosis, calcification, cysts, and bleeding on each section with reference to T1WI, T2WI, and DWI images. Additionally, an ROI section was drawn for the normal myometrium. Subsequently, the contrast ratio between the tumor and myometrium was calculated using the T1WI and T2WI images.

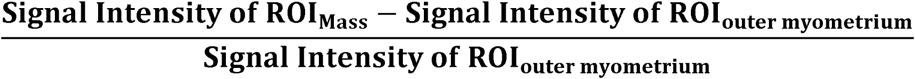

For masses with multiple areas of enhancement, the analysis focused on the region that exhibited the highest degree of enhancement. To determine whether the lesion was restricted, an ROI was placed over the solid portion of the tumor on each section of the ADC map. The ADC map was generated on a pixel-by-pixel basis, and the overall signal intensity of the DW images for a b-value of 1000s/mm2 was evaluated. The goal was to assess whether the lesion demonstrated high signal intensity on DW images and low signal intensity on the ADC map.

### Histopathologic examination

All histopathological specimens obtained during surgery underwent review by a gynecological pathologist. The final diagnosis regarding the origin of uterine carcinoma was established based on the histopathological findings, taking into account the clinical information and morphological features of the lesion as observed in MR images.

### Statistical analysis

Quantitative variables were presented using the mean and standard deviation (SD), while qualitative variables were presented using numbers and percentages (%). To compare data between groups, chi-square tests were used for categorical variables and independent samples t-tests were used for continuous variables. Receiver operating characteristic (ROC) analyses were performed to assess the ability of MRI parameters to distinguish between cervical and endometrial cancer or different cervical cancer pathologies (SCC or Adenocarcinoma). The overall performance of the ROC analysis was quantified by calculating the area under the curve (AUC), and optimal cut-off values, sensitivity, and specificity values were determined for significant parameters. Each variable was examined independently in a linear regression model using logistic stepwise regression. Data analysis was conducted using IBM SPSS Software, version 25.0. Two-tailed testing was employed, and p-values < 0.05 were considered statistically significant.

## Results

In our study, a total of 78 patients with uterine carcinoma were enrolled, with ages ranging from 20 to 80 years. The distribution of the types of uterine carcinoma in the study population was as follows: 20 out of 78 had endometrial cancer, 21 out of 78 had SCC of the cervix, 15 out of 78 had adenocarcinoma of the cervix, and 22 out of 78 had uterine carcinoma involving both the cervix and endometrium, with an indistinct origin based on initial biopsy and clinical findings.

Upon histological examination, the final diagnosis was confirmed as follows: 20 out of 78 were diagnosed with adenocarcinoma of the cervix, 27 out of 78 were diagnosed with SCC of the cervix, and 31 out of 78 were diagnosed with endometrial adenocarcinoma.

The location of the tumor was found to be statistically significant in determining the origin of the tumor. Our analysis revealed that 76.6% of cervical cancers and 64.5% of endometrial cancers were located accordingly, supporting the association between tumor location and origin.

In addition, certain imaging features were found to be consistent with cervical carcinoma. These included parametrial invasion (69.6%), vaginal invasion (66%), stromal invasion (95.7%), and peripheral rim enhancement (68.9%). These features significantly differed from those observed in endometrial carcinoma (p-value < 0.001). (figure 1)

**Figure 1.**
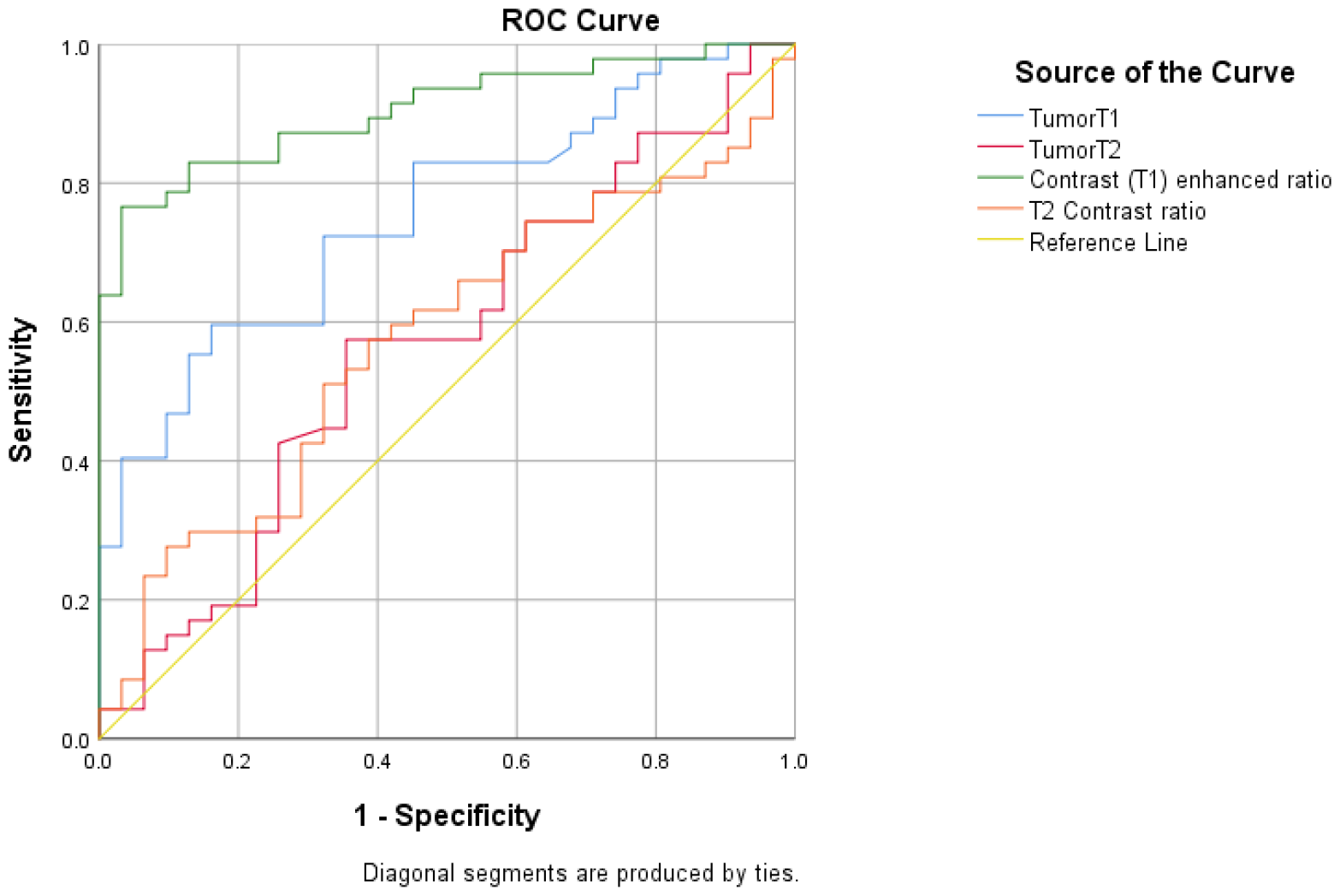
ROC curve for the discriminant ability of quantitative MRI features

**Figure 2.**
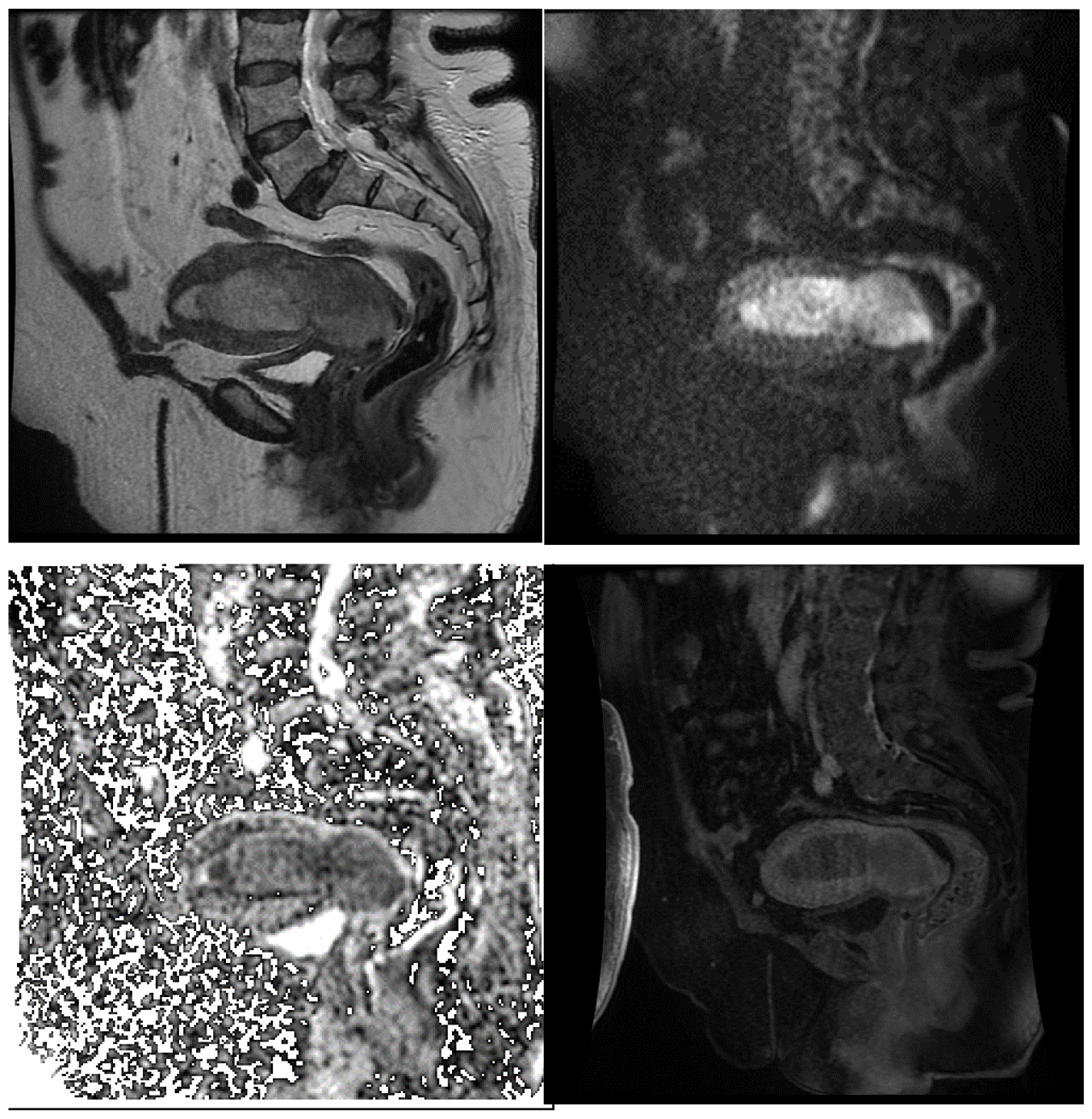
A lady in her 60s with cervical adenocarcinoma. Note that the mass shows intermediate signal on T2WI, with restricted diffusion on DWI, low ADC and enhancement less than myometrium.

On post-contrast sequences, cervical cancer appeared hyperintense compared to the myometrium, while endometrial cancer appeared hypointense (96.8%). This difference was statistically significant (p-value < 0.001). However, the presence of fluid retained in the distended endometrial cavity was not able to discriminate between the origins of the tumors, as there was no significant difference between cervical and endometrial tumors in this specific feature (Table 1).

**Table 1.** Incidence of MRI features in endometrial and cervical cancers.

|  | **Cervical**<br>**cancer**<br>**n=47** | **Endometrial**<br>**cancer**<br>**n=31** | **Odd ratio** | **p-**<br>**Value** |
| --- | --- | --- | --- | --- |
| Pathology |  |  |  |  |
| • Adenocarcinoma | 20 (42.6%) | 31 (100.0%) | 0.000 | <0.001 |
| • SCC | 27 (57.4%) | 0 (0.0%) |  |  |
| Site of tumor |  |  |  |  |
| • Cervix | 36 (76.6%) | 0 (0.0%) | - | <0.001 |
| • Endometrium | 0 (0.0%) | 20 (64.5%) |  |  |

|  | **Cervical cancer**<br>**n=47** | **Endometrial cancer**<br>**n=31** | **Odd ratio** | **p-Value** |
| --- | --- | --- | --- | --- |
| • Both | 11 (23.4%) | 11 (35.5%) |  |  |
| Peripheral rim enhancement |  |  |  |  |
| • Yes | 31 (68.9%) | 1 (3.2%) | 0.015 | <0.001 |
| • No | 14 (31.1%) | 30 (96.8%) |  |  |
| Mass in endometrial cavity |  |  | - | <0.001 |
| • Yes | 10 (21.7%) | 31 (100.0%) |  |  |
| • No | 36 (78.3%) | 0 (0.0%) |  |  |
| Depth of myometrium invasion > 50% |  |  | 13.054 | <0.001 |
| • Yes | 4 (8.5%) | 17 (54.8%) |  |  |
| • No | 43 (91.5%) | 14 (45.2%) |  |  |
| Distended filled endometrial cavity |  |  | 0.928 | 0.892 |
| • Yes | 11 (23.9%) | 7 (22.6%) |  |  |
| • No | 35 (76.1%) | 24 (77.4%) |  |  |
| Parametrical invasion |  |  | 0.067 | <0.001 |
| • Yes | 32 (69.6%) | 4 (13.3%) |  |  |
| • No | 14 (30.4%) | 26 (86.7%) |  |  |
| Vaginal invasion |  |  | 0.055 | <0.001 |
| • Yes | 31 (66.0%) | 3 (9.7%) |  |  |
| • No | 16 (34.0%) | 28 (90.3%) |  |  |
| Enhancement related to myometrium |  |  | 0.021 | <0.001 |
| • Hypo | 18 (38.3%) | 30 (96.8%) |  |  |
| • Hyper | 29 (61.7%) | 1 (3.2%) |  |  |
| Craniocaudal and Transverse |  |  | 0.000 | <0.001 |
| • C ≥ T | 18 (38.3%) | 31 (100.0%) |  |  |
| • C < T | 29 (61.7%) | 0 (0.0%) |  |  |
| Stromal invasion |  |  | 0.015 | <0.001 |
| • Yes | 45 (95.7%) | 8 (25.8%) |  |  |
| • No | 2 (4.3%) | 23 (74.2%) |  |  |

Furthermore, endometrial carcinoma was well diagnosed by the presence of an endometrial cavity mass (100%), deep myometrium invasion (>50%) (54.8%), and a greater size in the craniocaudal dimension compared to the transverse dimension (100%).

The analysis of quantitative parameters revealed no significant difference between cervical and endometrial cancers in terms of mean ADC value, tumor size, and signal intensity of T2-weighted images, T2 ratio, and T2 contrast ratio. However, there were significant differences between cervical and endometrial carcinoma in terms of the signal intensity of T1-weighted images (765.77 for cervical carcinoma, 514.48 for endometrial carcinoma), T1 ratio (1.09 for cervical carcinoma, 0.61 for endometrial carcinoma), and T1 contrast enhancement ratio (0.09 for cervical carcinoma, −0.39 for endometrial carcinoma) (p-value < 0.001) (Table 2).

**Table 2.** Incidence of quantitative MRI features in endometrial and cervical cancers.

|  | **Cervical cancer** | **Endometrial cancer** | **p-Value** |
| --- | --- | --- | --- |
| Mean adc | 802.30±149.70 | 786.97±125.87 | 0.640 |
| Size (mm) | 52.26±21.56 | 54.34±20.97 | 0.682 |
| Tumor T1 | 765.77±273.71 | 514.48±202.40 | <0.001 |
| Tumor T2 | 612.79±643.06 | 492.74±170.54 | 0.314 |
| T1 ratio* | 1.09±0.36 | 0.61±0.12 | <0.001 |
| T2 ratio* | 1.76±1.39 | 1.47±0.38 | 0.253 |
| T1 contrast ratio** | 0.09±0.36 | -0.39±0.12 | <0.001 |
| T2 contrast ratio** | 0.76±1.39 | 0.47±0.38 | 0.253 |

ROC curves were used to assess the sensitivity and specificity of the tests. Figure 1 shows that the signal intensity of T1 and T1 contrast enhancement ratio had larger areas under the curves, indicating that they are more accurate diagnostic tests. The AUC values were 0.755 for T1 and 0.905 for T1 contrast enhancement ratio, further supporting their diagnostic value in differentiating endometrial from cervical cancers (Table 3).

**Table 3.** Diagnostic value of quantitative parameters to distinct cervical cancers from endometrial cancers.

| Test Result Variable(s) | AUC* | p-Value | 95% Confidence Interval |  |
| --- | --- | --- | --- | --- |
|  |  |  | Lower Bound | Upper Bound |
| TumorT1 | 0.755 | <0.001 | 0.649 | 0.860 |
| TumorT2 | 0.568 | 0.315 | 0.436 | 0.699 |
| Contrast (T1) enhanced ratio | 0.905 | <0.001 | 0.840 | 0.970 |
| T2 Contrast ratio | 0.575 | 0.264 | 0.447 | 0.703 |

\*AUC: area under curve

The optimal cutoff for the signal intensity of T1 in discriminating the origin of uterine tumors was estimated to be 592. At this cutoff, the sensitivity was 72.3%, specificity was 67.7%, negative predictive value (NPV) was 61.8%, and positive predictive value (PPV) was 77.3%.

For the T1 contrast enhancement ratio, the cutoff point was −0.25, with a sensitivity of 83%, specificity of 87.1%, NPV of 77.1%, and PPV of 90.7%. These values indicate the ability of the T1 contrast enhancement ratio to accurately differentiate between cervical and endometrial cancers (Table 4).

**Table 4.** Predictive value and cut-off point for discriminant quantitative MRI features.

| Test Result Variable(s) | Overall Accuracy (%) | AUC | Cut off | Sen<sup>1</sup> (%) | Spec<sup>2</sup> (%) | NPV<sup>3</sup> (%) | PPV<sup>4</sup> (%) | BER<sup>5</sup> (%) |
| --- | --- | --- | --- | --- | --- | --- | --- | --- |
| TumorT1 | 70.5 | 0.755 | 592 | 72.3 | 67.7 | 61.8 | 77.3 | 30 |
| Contrast (T1) enhanced ratio | 84.6 | 0.905 | -0.25 | 83 | 87.1 | 77.1 | 90.7 | 15 |

1: Sensitivity, 2: Specificity, 3: Negative Predictive Value, 4: Positive Predictive Value, 5: Balanced Error Rate

Stepwise regression was performed by adding one independent variable at a time to examine the statistical significance of each variable in the regression model. Forward stepwise regression analysis revealed that the following variables were significant in differentiating endometrial and cervical cancers as independent variables:

1. Deep myometrium invasion >50% (odds ratio 0.029)
2. Stromal invasion (odds ratio 48.870)
3. Vaginal invasion (odds ratio 262.388)
4. Contrast (T1) enhanced ratio > −0.25 (odds ratio 20.257)

These variables showed statistical significance and their odds ratios indicate their association with the differentiation between endometrial and cervical cancers (Table 5).

**Table 5.** Stepwise regression for discriminant MRI features. OR: odds ratio, CI: confidence interval.

|  | OR | 95% CI |  | P-value |
| --- | --- | --- | --- | --- |
| Depth of myometrium invasion > 50% | 0.029 | 0.001 | 0.617 | 0.023 |
| Vaginal invasion | 48.870 | 2.172 | 1145.057 | 0.014 |
| Stromal invasion | 262.388 | 5.073 | 13571.027 | 0.006 |
| Contrast (T1) enhanced ratio >-0.25 | 20.257 | 1.267 | 323.806 | 0.033 |

## Discussion

The diagnosis of uterine carcinoma, specifically distinguishing between endometrial and cervical cancers, is primarily done through physical examination and tumor biopsy. However, there are challenges in diagnosing a small subset of tumors with an indistinct origin or those that involve both the cervix and endometrium. This poses difficulties in determining the appropriate treatment for these cases (11,12,13). The FIGO staging system, which is commonly used for uterine tumors, does not provide clear guidelines for tumors that extend and involve both the cervix and endometrium. As a result, staging and treatment for these tumors remain unclear (14). MRI is often the preferred modality for studying tumor morphology and features. Some studies have assessed MRI features to differentiate between cervical and endometrial carcinomas. Vergas et al. reported an overall accuracy of MRI in detecting the origin of uterine carcinoma to be 85%-88% in a retrospective study (3). Haidar et al. identified outstanding MRI features such as endometrial thickening or mass expanding the endometrial cavity and tumor invasion of the myometrium, which can help differentiate endometrial adenocarcinoma from cervical adenocarcinoma (15). Previous studies have also estimated the overall accuracy of MRI for staging endometrial and cervical carcinomas to range from 85% to 93% and 75% to 96%, respectively (16,17,18). The current prospective study aimed to describe morphologic features that have promising power in discriminating between cervical and endometrial carcinoma.

The study found that certain morphologic features were reliable indicators for detecting cervical carcinoma, including vaginal, stromal, and parametrial invasions. Additionally, the presence of hypervascularity in the tumor using the myometrium as an internal reference and peripheral rim enhancement increased the likelihood of cervical cancer. On the other hand, myometrial invasion and the presence of a mass in the endometrial cavity were significantly higher in endometrial carcinoma. These findings align with a previous study by Bourgioti et al., who also reported MRI features that could distinguish cervical carcinoma from endometrial carcinoma. These features included tumor location, enhancement on early arterial dynamic contrast-enhanced MRI (DCE-MRI), peripheral enhancing rim, mass in the endometrial cavity, myometrial invasion >50%, and full-depth cervical stromal invasion. However, the current study did not assess features such as adnexal invasion, pelvic side wall/adjacent organ involvement, and enhancement on late DCE-MRI, which were considered invaluable for discriminating the origin of the tumor. Furthermore, the current study introduced the craniocaudal dimension as a variable, which had not been previously assessed. This variable was found to be capable of distinguishing endometrial carcinoma from cervical carcinoma (11). Our analysis of quantitative MRI features showed that signal intensity of T1WI, T1 ratio and contrast enhanced (T1) ratio were useful to evaluate the origin of tumor. In addition, signal intensity of T1 and T1 contrast-enhanced ratio with AUC equal to 0.755 and 0.905, respectively, had higher diagnostic value to determine the origin of the tumor.

Our study found that the ADC was not a reliable variable for distinguishing the origin of tumors. However, Lin et al. discovered that the mean ADC value was significantly higher in cervical carcinoma compared to endometrial carcinoma. This difference could be attributed to variations in the techniques used for measurement and description of the ADC value. In their study, Lin et al. developed a MDS (multidimensional scoring) system that combined quantitative DW MRI, MR spectroscopy, and morphological features of MR imaging. In contrast, our study did not utilize a scoring system. Instead, we aimed to determine the diagnostic value of each parameter independently in discriminating the origin of uterine carcinoma. Our stepwise regression analysis revealed that myometrial invasion, stromal invasion, vaginal invasion, and the contrast T1 enhanced ratio were significantly higher in favor of distinguishing cervical carcinoma from endometrial carcinoma.

Like any study, our current study has several limitations that should be considered. Firstly, we did not establish the kappa coefficient, which resulted in an undetermined level of agreement for the imaging criteria used in our study. Furthermore, it is important to note that our study did not include patients with microscopic disease or rare types of carcinoma, such as serous and clear cell carcinomas. Additionally, the sample size in our study was limited, which may affect the generalizability of the findings. Another limitation of our study is that we did not consider the time and cost effectiveness of the diagnostic methods used. Future research should take into account these factors in order to provide a comprehensive evaluation of the diagnostic approach.

To further advance our understanding, it is crucial to continue investigating with a larger sample size that includes patients with tumors involving both the endometrium and cervix. Additionally, it would be beneficial to determine the kappa value to establish the level of agreement for the imaging criteria used in the study.

## Conclusion

Indeed, MRI has become increasingly valuable in detecting and differentiating between cervical and endometrial cancers. Various MR features have been identified as useful in discriminating between these two types of cancers. These features include tumor location, early arterial enhancement, peripheral rim enhancement, parametrial invasion, vaginal invasion, stromal invasion, myometrium invasion, endometrial cavity mass, craniocaudal dimension, signal intensity of T1, T1 ratio, and contrast-enhanced T1 ratio.

By developing a multiparameter model based on these MRI features, radiologists can enhance their ability to accurately diagnose the origin of the tumor. This approach can significantly improve diagnostic accuracy and aid in appropriate treatment planning and management for patients with uterine cancers.

## Data Availability

All data produced in the present work are contained in the manuscript

## References

1. Siegel RL, Miller KD, Jemal A. Cancer statistics, 2017. CA Cancer J Clin 2017; 67:7–30.

2. NCCN Clinical Practice Guidelines in Oncology: Uterine Cancers. National Comprehensive Cancer Network Web site. http://www.nccn.org/professionals/physician_gls/pdf/uterine.pdf. Accessed August 22,2017.

3. Vargas, Hebert Alberto, et al. “The value of MR imaging when the site of uterine cancer origin is uncertain.” Radiology 258.3 (2011): 785.

4. Colombo N, Carinelli S, Colombo A, et al. (2012) Cervical cancer: ESMO Clinical Practice Guidelines for diagnosis, treatment andfollow-up. Ann Oncol 23(Suppl 7):vii27–32

5. Colombo N, Preti E, Landoni F, et al. (2013) Endometrial cancer: ESMO Clinical Practice Guidelines for diagnosis, treatment and follow-up. Ann Oncol 24(Suppl 6):vi33–38

6. Freeman SJ, Aly AM, Kataoka MY, Addley HC, Reinhold C, Sala E. The revised FIGO staging system for uterine malignancies: implications for MR imaging. Radiographics. 2012;32(6):1805–27.

7. Otero-García, María Milagros, et al. “Role of MRI in staging and follow-up of endometrial and cervical cancer: pitfalls and mimickers.” Insights into imaging 10.1 (2019): 1–22.

8. Lee SI, Atri M. 2018 FIGO staging system for uterine cervical cancer: enter cross-sectional imaging. Radiology. 2019;292(1):15–24.

9. Hardesty LA, Sumkin JH, Nath ME, Edwards RP, Price FV, Chang TS, et al. Use of preoperative MR imaging in the management of endometrial carcinoma: cost analysis. Radiology. 2000;215(1):45–9.

10. Lin CN, Liao YS, Chen WC, Wang YS, Lee LW. Use of myometrium as an internal reference for endometrial and cervical cancer on multiphase contrast-enhanced MRI. PLoS One 2016;11: e015782

11. Bourgioti, Charis, et al. “Endometrial vs. cervical cancer: development and pilot testing of a magnetic resonance imaging (MRI) scoring system for predicting tumor origin of uterine carcinomas of indeterminate histology.” Abdominal imaging 40.7 (2015): 2529–2540.

12. McCluggage WG. Ten problematical issues identified by pathology review for multidisciplinary gynaecological oncology meetings. Journal of clinical pathology. 2012;65(4):293–301.

13. He H, Bhosale P, Wei W, Ramalingam P, Iyer R. MRI is highly specific in determining primary cervical versus endometrial cancer when biopsy results are inconclusive. Clin Radiol 2013; 68:1107–1113.

14. Salvo, Gloria, et al. “Revised 2018 International Federation of Gynecology and Obstetrics (FIGO) cervical cancer staging: a review of gaps and questions that remain.” International Journal of Gynecologic Cancer 30.6 (2020).

15. Haider, Masoom A., et al. “Adenocarcinoma involving the uterine cervix: magnetic resonance imaging findings in tumours of endometrial, compared with cervical, origin.” JOURNAL-CANADIAN ASSOCIATION OF RADIOLOGISTS 57.1 (2006): 43.

16. Ramirez PT, Frumovitz M, Milam MR, et al. (2010) Limited utility of magnetic resonance imaging in determining the primary site of disease in patients with inconclusive endometrial biopsy. Int JGynecol Cancer 20:1344–1349

